# Workplace Violence and Self-Reported Physical and Mental Health: A National Cross-Sectional Study in Lebanon

**DOI:** 10.1101/2025.04.05.25325303

**Authors:** Hazar Shamas, Ghada E. Saad, Myriam Dagher, Rita Itani, Ali Abboud, Stephen J. McCall, WOMENA Study Group

## Abstract

**Background:** Workplace violence (WPV) is any physical or psychological violence experienced in the workplace. Studies that investigate the prevalence of WPV in the Middle East North Africa (MENA) region are scarce. This study aimed to examine the determinants of WPV and its association with self-reported physical and mental health among employed adults residing in Lebanon.

**Methods:** This was a national cross-sectional study that recruited working age residents of Lebanon through Random Digital Dialing. Data were collected by trained data collectors through SurveyCTO software from January till July 2024. The main exposure of this study was physical and verbal WPV. Three outcomes depressive symptoms, anxiety symptoms, and physical health were measured using the PHQ-9, GAD-7, and PROMIS GBH scales, respectively. Adjusted logistic regression models estimated the association between WPV and each outcome, respectively.

**Findings:** The study included 3,076 employed participants. Median age (IQR) of study sample was 37 (28-46) years old, 889 (30%) completed college and further education, 1,111 (25%) were non-Lebanese. A total of 518 (16%) participants experienced at least one form of WPV. Moreover, 1,080 (33·4%) experienced depressive symptoms, 825 (25·5%) experienced anxiety symptoms, and 2,374 (75·5%) reported poor physical health. Being exposed to WPV increased the odds of depressive symptoms [adjusted odds ratio (aOR):3·00 (95%CI:2·40-3·70)], anxiety symptoms (aOR:3·01 (95%CI:2·41-3·72)), and poor physical health (aOR:2·82 (95%CI:2·04-3·98)).

**Interpretation:** The study highlights the extent of WPV among workers in Lebanon and the urgent need to address the matter. Findings offer a basis for targeting interventions to vulnerable workers.

**Funding:** International Development Research Centre (IDRC)- Canada

**Research in context:** *Evidence before this study:* A literature search was conducted on workplace violence (WPV) exposure and mental and physical health outcomes using the following databases: PubMed, Web of Science, Science Direct, and Google Scholar. Articles published in English until September 1, 2024 were searched using the search terms “workplace violence”, “occupational violence”, “workplace physical violence”, “workplace sexual violence”, “workplace sexual harassment”, and “mental health”, “depression”, “anxiety”, “trauma”, “isolation”, “social phobia”, “physical health”, “non-communicable diseases, “cancer”, “cardiovascular disease”, “diabetes”, “hypertension”, “respiratory disease”, “anemia”, “musculoskeletal disease”, “chronic renal failure”, “neurological disease”, “sleep problems”, “sick leave”, “sickness”, and “population-based study”. The Lancet 2023 Series “Work and Health” highlighted the elevated risk of experiencing anxiety symptoms, depressive symptoms, and overall fatigue for workers experiencing WPV. The series accentuated the scarcity of studies examining the impact of WPV on mental and physical health in low-middle income countries (LMIC). Studies that aim to investigate the magnitude of WPV in Lebanon and in the Middle East and North Africa (MENA) region were only conducted within the healthcare sector focusing on healthcare workers.

*Added value of this study:* This study adds significant value as one of the largest national studies in the MENA region to examine the association of WPV with mental and physical health among adults in all work sectors. This is the first study that investigates association of WPV with depressive symptoms, anxiety symptoms, and self-reported physical health among Lebanese and non-Lebanese working in the formal and informal sector and residing in Lebanon, a LMIC. Results reveal a relatively high prevalence of WPV in both work sectors. Study results also show a significantly higher odds of WPV among uneducated, non-Lebanese including refugees and immigrants, and freelance informal sector workers. Furthermore, WPV was strongly associated with depressive symptoms, anxiety symptoms, and reporting poor physical health.

*Implications of all the available evidence:* This large national study highlights the adverse mental and physical health outcomes of WPV on employed adults in Lebanon particularly among non-nationals including immigrants and refugees, uneducated, and informal freelance workers. These findings accentuate detrimental health, social, and financial implications of WPV on institutions and organisations. The findings suggest potential benefit from enhancing labour law enforcement by the government and providing targeted interventions that promote respect and tolerance within formal and informal work sectors regardless of worker’s nationality.

## Introduction

Workplace violence (WPV) is defined as any physical or psychological violence experienced in the workplace.^1,2^ Workplace physical violence involves a range of actions including sexual assault, hitting, spitting, shoving, and use of weapons.^1,2^ Whereas workplace psychological violence manifests as verbal threats, bullying, manipulation, micromanagement, and isolation.^1,2^ WPV is a form of stigmatisation, discrimination, and inequality.^2^ Globally, almost 23% of employed adults experience at least one form of violence and harassment at work of which 18% are psychological.^3^ WPV can have long-term financial effects on the organisation where the Lancet 2023 series “Work and Health” emphasised the influence of WPV on reduced productivity and increased turnover.^4^ This series also highlighted the elevated risk of experiencing anxiety symptoms, depressive symptoms, and overall fatigue for workers enduring WPV.^4,5^

Lebanon is a low-middle income country (LMIC) situated in the Middle East and North Africa (MENA) region. The country has been combating repercussions of the Beirut port blast, political instability, and economic crisis. These crises have led to a significant portion of the population enduring financial hardship.^6^ In addition, Lebanon also hosts the highest number of refugees per capita in the world, where a quarter of the population are refugees, most of which are Syrians who lack residency permits and work without written contracts.^7,8^ Studies have shown that a significant share of Lebanon’s labour market relies on Syrian refugees, many of whom are discriminated against, informally employed, have no legal protection, and are subjected to workplace exploitation.^4,9,10^

Globally, studies have shown that WPV is associated with increased risk of sickness absence, sleep-related problems, stress, burnout, binge drinking, social phobia, suicide attempts, suicide, poor physical and mental health.^11-15^ However, most studies were conducted in high-income countries and targeted the formal sector, particularly those who work in the healthcare sector.^11-14^ Studies in the MENA region also highlighted the impact WPV has on deteriorating healthcare workers’ mental and physical health.^16-18^ All studies reporting the proportion of WPV in the work setting indicated that psychological violence was more prevalent than physical violence at work.^14,16-18^ Similarly, in Lebanon, all studies that investigated the impact of WPV were limited to the healthcare sector and focused on healthcare workers, doctors, and nurses.^19,20^ Findings from those studies revealed a high prevalence of WPV among nurses across Lebanon, where 62% reported verbal abuse and 10% reported physical violence.^19,20^ A comprehensive understanding of the impact of WPV on health in all sectors of employment is needed in low resource settings; particularly, especially in settings that have a high proportion of informal labour and refugee populations. Thus, the aim of this study was to examine the determinants of WPV and its association with self-reported physical and mental health among Lebanese and non-Lebanese employed adults residing in Lebanon.

## Methods

### Study design and setting

This was a national cross-sectional telephone survey in Lebanon. This survey formed part of a larger project titled: “Identifying opportunities to improve the lived experience and health of working women in the MENA: from COVID to recovery-WOMENA”.

### Sampling and study population

Residents in Lebanon were recruited through Random Digit Dialing (RDD) between January and July 2024. Numbers dialed were constructed within a 11-digits frame used by mobile phones in Lebanon. The first 3 numbers constituted of +961, the international country calling code for Lebanon. The code is usually succeeded by two digits implying the country’s mobile network operators (i.e., 03, 70, 71, 76, 78, 79, 81). The remaining 6 numbers of the 11 were randomly generated. Contact with each generated phone number was attempted twice, in case there was no answer the first time. If respondents were unable to complete the survey, follow-up calls were scheduled for a convenient time. To reduce reporting bias, calls were made either during weekdays after working hours or on Saturdays between 09:00 and 19:00 Beirut local time.

Respondents had to answer a group of screening questions. Study eligibility criteria included residents in Lebanon who are employment age. Women were oversampled to ensure sufficient number of employed men and women. This current study population was restricted to those who were employed.

Informed oral consent was obtained from all participants prior to their involvement in the study. A strategic monitoring plan with standardised data quality procedures was adopted while collecting data. Data was monitored weekly to detect and mitigate systematic errors and missing data, in a timely manner. Five percent of surveys were recorded for quality checking purposes. The recordings were compared with the collected and entered data to identify any data errors.

The error rate, computed as the number of identified errors out of the total number of answered questions, was calculated and returned a 0·59% error rate. Identified errors in data entry were corrected with the help of the recordings or, if necessary, follow-up calls with participants whose data was incorrectly entered.

### Data sources

The study survey incorporated existing questionnaire modules, scales, and community-identified priorities adapted to the Lebanese context. The survey was initially developed in English then translated to Arabic. Data were collected by trained data collectors in both languages through SurveyCTO software. Participants who consented and were eligible for participation had to answer questions regarding their socio-demographics, employment history, physical fitness, anxiety and depressive symptoms.

### Main exposure

The main exposure of this study was WPV. WPV was defined using the following question extracted from the International Labour Organization Gallup (ILO): during the past 6 months, have you ever experienced at least one of the following a) physical violence and/or harassment at work, such as hitting, restraining, or spitting b) psychological violence and/or harassment at work, such as insults, threats, bullying, or intimidation?^21^

### Outcomes of interest

Three outcomes were explored in this study: depressive symptoms, anxiety symptoms, and self-reported physical health. Depressive symptoms were measured through the Patient Health Questionnaire-9 (PHQ-9).^22^ PHQ-9 is a 9-item questionnaire assessing depressive symptoms. Its score ranges from 0 to 27 where each item is scored from 0 (not at all) to 3 (nearly every day).^22^ A total score of 10 or more on the PHQ-9 indicated the presence of depressive symptoms.^22^

Anxiety symptoms were measured through the General Anxiety Disorder-7 (GAD-7).^23^ GAD-7 is a 7-item questionnaire that assesses anxiety symptoms. Its score ranges from 0 to 21 where each item has a score of 0 (not at all) to 3 (nearly every day). A total score of 10 or more on the GAD-7 indicated the presence of anxiety symptoms. Thus, this cut-off point was used for this study.^23^

Physical health was measured using the PROMIS GBH 1·2.^24^ This scale included 4 questions indicating physical health. Score for each question ranges from 1 to 5 where 1 indicated poor physical health and 5 excellent physical health. Subsequently, each questions’ scores were summed up and dichotomised into good physical health, defined as the highest 25^th^ percentile of scores and poor physical health.^24^

The PHQ-9, GAD-7, and PROMIS GBH scale had excellent reliability in this population with a Cronbach’s alpha of 0·80, 0·85, and 0·80 respectively.

### Confounders

Confounders of this study were identified using the Directed Acyclic Graphs (DAGs).^25^ DAGs were constructed based on causal paths between the main exposure (WPV), and the three outcomes depressive symptoms, anxiety symptoms, and physical health (Figures S1, S2, and S3 in Appendix p 3 and 4). The confounding pathways of the association between WPV and the three outcomes were age, sex, education, nationality, marital status, urbanicity, and job sector.

Urbanicity was assessed using the single-item self-report measure (SIDU): “please indicate how urban your living environment is on a 7-point scale from 1 indicating not urban at all to 7 meaning very urban.” The responses were then dichotomised where a score of 6 and more implying extremely urbanised.^26^ Participants were attributed to one job sector depending on their work. The job sectors included government/ NGO worker, private business worker, private household worker, and freelance/ informal sector worker.

### Statistical analysis

To account for the study design that aimed to have equal allocation of employed and unemployed males and females, sampling and post calibration weights were calculated to allow for national estimates. Determinants of WPV as well as their association with the outcomes were presented and identified through the literature. Frequency of participants reporting each outcome along with their weighted percentages, unadjusted weighted odds ratios (ORs) and 95% confidence intervals (CIs) were calculated. Variables with p-values less than 0·05 were considered statistically significant. Separate logistic regressions were run to estimate the association between workplace violence, depressive and anxiety symptoms, and physical health.

Missing values were tested using Little’s test of missing completely at random (MCAR). The maximum number of missing values in the variables was 1·5% which was MCAR. Therefore, complete case analysis was implemented.^27^

### Ethical and committee approvals and reporting

This study followed the Strengthening the Reporting of Observational Studies in Epidemiology (STROBE) guidelines for reporting.^28^ The study protocol was reviewed and approved by the American University of Beirut Social and Behavioral Sciences Institutional Review Board during November 2023 [Reference: SBS-2023-0182]. All procedures performed followed the institution’s ethical guidelines including informed consent, confidentiality, and the right to withdraw from the study at any point.

## Results

A total of 97,608 calls were placed using RDD. Of those calls, 32,411 (33%) responded while the rest were unreachable. Among those who responded, 73% did not consent, and 0.03% were previously contacted by the team on their second phone number. Of the 8,599 that consented to be part of the study, 14% were either below or above the eligible age range of 19-64 years old. Therefore, a total of 4,725 were eligible and agreed to participate out of which 3,076 were employed. Thus, this study included 3,076 participants (Figure 1). The frequency of participants having each outcome along with their weighted percentages, unadjusted weighted odds ratios were presented in Appendix p 5-7.

**Figure 1.**
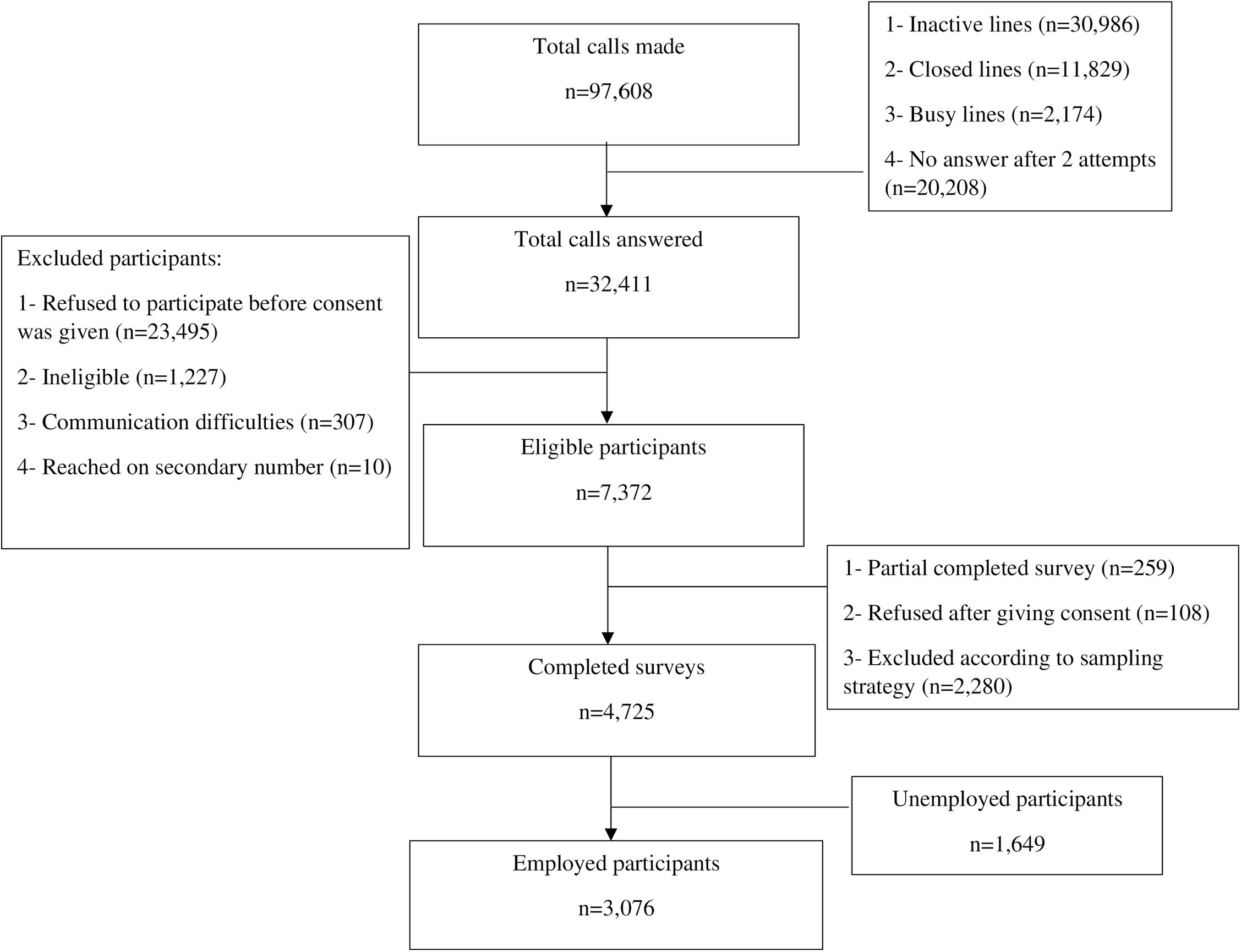
Flow diagram representing the participants included in the study

The median age (IQR) of the study participants was 37 (28-46) years old, 889 (30%) completed college and further education, 2,130 (68%) were married or engaged and 1,111 (25%) were non-Lebanese. A total of 518 (16%) participants experienced at least one form of WPV. Of those 518 participants, 36 (7%) experienced physical violence, 374 (72·2%) participants experienced psychological violence, and 108 (20·8%) participants experienced both violence forms. Psychological violence was the most prevalent form of WPV experienced among participants (n=482 (93% of 518)).

Females had lower odds of experiencing WPV compared to males OR:0·74 (95%CI: 0·60-0·91). Similarly, those with intermediate/ technical education and those who completed college or further education had lower odds of experiencing WPV compared to those with no/ primary education OR:0·53 (95%CI: 0·42-0·67) and OR:0·39 (95%CI: 0·30-0·51), respectively. The odds of experiencing WPV was higher among non-Lebanese compared to Lebanese OR:3·01 (95%CI: 2·44-3·66). WPV was also higher among freelance workers compared to those who work in the government and non-governmental organisations (NGO) OR:2·04 (95%CI:1·42-2·92) and compared to those who work in private business OR:1.50 (95%CI:1.21-1.86) (Figure 2).

**Figure 2.**
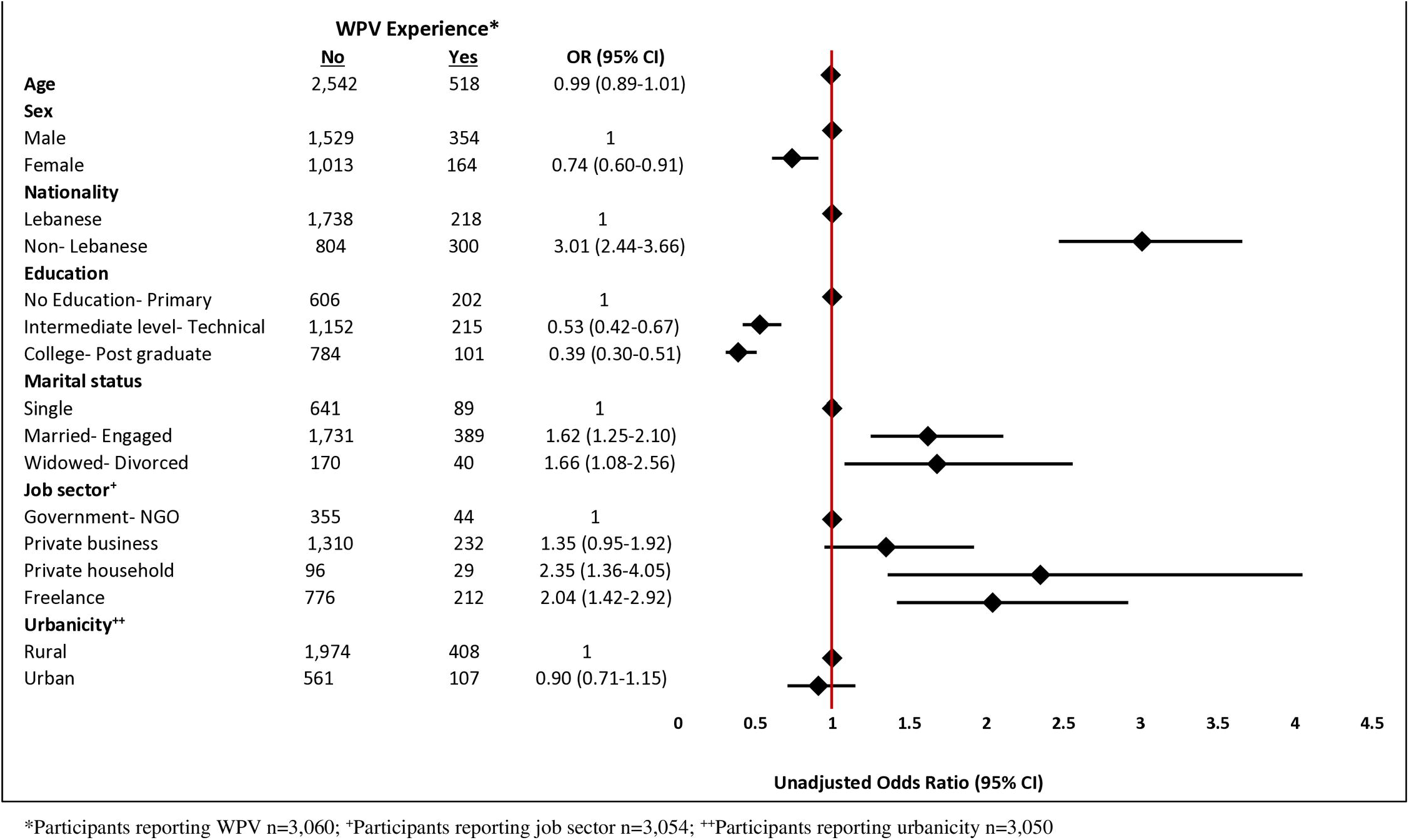
Unadjusted odds of workplace violence among socio-demographics

The odds of depressive symptoms were higher among non-Lebanese OR:1·99 (95%CI: 1·70-2·32), those who are widowed/ divorced OR:1·92 (95%CI: 1·38-2·67) and those who work in either private household OR:2·21 (95%CI: 1·37-3·27) or freelance OR:1·56 (95%CI: 1·20-2·02). The odds of depressive symptoms were lower among those who completed college or further OR:0·43 (95%CI: 0·35-0·53). Similarly, the odds of anxiety symptoms were higher among non-Lebanese OR:1·60 (95%CI:1·36-1·89), widowed/ divorced OR:1·93 (95%CI: 1·36-2·74), and those working in freelance OR:1·41 (95%CI: 1·07-1·84). Workers who are females, non-Lebanese, widowed/ divorced, and freelance workers had higher odds of reporting poor physical health OR:2·27 (95%CI: 1·86-2·76), OR:1·68 (95%CI: 1·39-2·04), OR:2·67 (95%CI: 1·69-4·06), OR:1·65 (95%CI: 1·24-2·20), respectively (Appendix p 5-7).

Accounting for age, sex, education, nationality, marital status, urbanicity, and job sector, the odds of depressive symptoms, anxiety symptoms, and poor physical health were almost 3 times higher among those who experienced workplace violence compared to those who did not [adjusted odd ratio (aOR):3·00 (95%CI: 2·40-3·70)], aOR:3·01 (95%CI: 2·41-3·72), and aOR:2·82 (95%CI: 2·04-3·98) for depressive symptoms, anxiety symptoms, and poor physical health, respectively (Figure 3).

**Figure 3.**
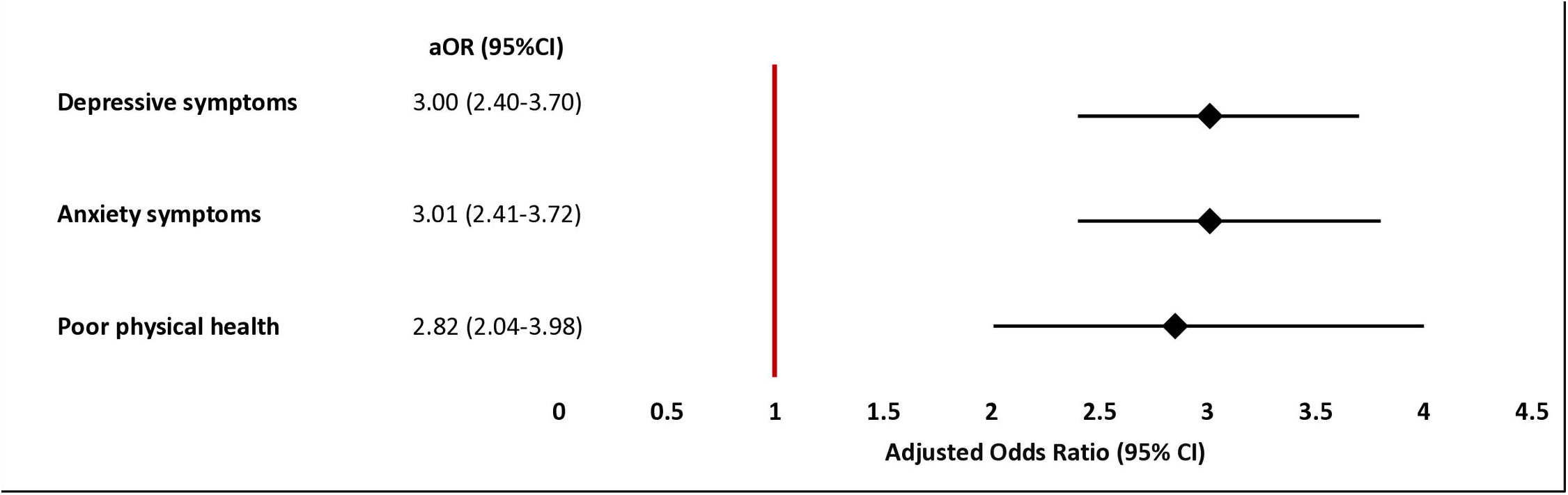
Adjusted odds of depressive symptoms, anxiety symptoms, and poor physical health

## Discussion

Nearly 1 of 6 employed adults in Lebanon experienced at least one form of WPV with psychological violence being the most common. WPV almost tripled the odds of displaying depressive symptoms, anxiety symptoms, and reporting poor physical health in a nationally representative sample of working-age adults in Lebanon. The odds of WPV were higher among males, uneducated, non-Lebanese residents, and freelance workers.

The prevalence of WPV among employed adults in Lebanon is relatively high compared to other countries.^14,29-31^ In 2020, Lebanon enacted law number 205 criminalising sexual harassment across different settings including the workplace regardless of nationality.^32^ However, this law is not strictly enforced in the workplace and workers must rely on the goodwill of their employer.^33^ Moreover, almost 31% of study participants belong to the freelance informal sector. Freelance workers are more prone to encounter WPV as they lack protection and formal support systems such as human resources department and compensation insurance.^4,10,34^ In addition, freelance workers are likely to have volatile job circumstances and are less likely to report or address incidents of WPV due to fear of losing income or future work opportunities.^33,35^ Another explanation for the high prevalence WPV is the large proportion of non-nationals in Lebanon. WPV has been shown to be higher among non-national workers of whom are mostly refugees or migrant workers.^7,36^ These populations often lack a social network or legal protections to prevent discrimination in the workplace.^10,37^ In addition, many refugees in Lebanon lack legal residency permits and have oral contracts with their employers or no contracts.^8,10^ As such, refugees can only join informal work sectors, which lends itself to exploitation and poor working conditions.^10^

Similar to multiple systematic reviews, WPV was associated with lack of self-esteem, depressive symptoms, anxiety symptoms, and poor physical health.^12,13,38^ This may be explained by the social consequences of WPV that manifest in the form of social isolation, job dissatisfaction, burnout, and poor engagement leading to reduced productivity, lack of motivation, lower self-esteem, and poor mental health.^38^ The impact of WPV varies by individual but could be considered as a traumatic event that leads to the feelings of fear, shame, chronic stress, helplessness, and loss of control inducing depression and anxiety. WPV was also associated with poor physical health. This may be a result of physical injuries, pain, back pain, headache/eye strain, overall fatigue, and possible disability.^39,40^

The study had few limitations. This was a cross-sectional study where temporality was not met; therefore, it is essential to conduct follow up this population in the future. The outcomes of this study were self-reported, which could also increase misclassification bias. Moreover, data were collected through trained data collectors, and 5% of the recorded interviews were quality-checked with only a 0.5% error rate which limits information bias. This was one of the largest national cross-sectional studies in the MENA region examining the association of WPV with mental and physical health. Furthermore, there are very few nationally representative studies in the MENA region that explored the WPV amongst all work sectors, especially the informal sector which is at greatest risk. Additionally, this study adds to the literature by exploring the association between WPV and depressive and anxiety symptoms using reliable and validated scales.

Building on the gaps highlighted by the Lancet “Work and Health” series, we have shown the serious implications of experiencing WPV on employee’s physical and mental health. The results illustrate that there should be better efforts from the government to implement and enforce law 205 that ensures a safe, secure, and productive work environment for employees in the informal and formal work sectors.^5^ In addition, there is a requirement to improve the safeguards, whistleblowing, disciplinary actions and anti-violence policies in workplaces in Lebanon.^5,41^

WPV is a critical issue that affects workers’ health. This study sheds light on the extent of WPV among workers in Lebanon and the urgent need to address the matter. The study highlights the vulnerabilities of workers who are non-nationals, uneducated, and in unstable jobs, which increase the odds of WPV. Findings offer a basis for targeting interventions that promote respect and tolerance between workers within the workplace.

## Data Availability

Data is anonymised and can be shared upon reasonable request from the Center for Research on Population and Health at the American University of Beirut.

## Authors’ contributions

SJM, HG, GS and AA conceptualised the survey component of study. SJM, HG, AA, MD, and GS designed the survey, contributed to study methodology and investigation. MD oversaw the data collection and was supervised by SJM. RI secured ethical approval of the study. The formal analysis and literature search for this study were conducted by HS. HS and SJM drafted the manuscript. SJM supervised HS throughout the project. HS, SJM, HG, AA, MD, RI, and GS contributed to the interpretation of the results, edited drafts, and approved the final version of the article. HS, SJM, HG, AA, MD, RI, and GS had full access to and verified the raw data. All authors had access to the study data and had final responsibility for the decision to submit the manuscript for publication.

## Data sharing

### Acknowledgments

This work was funded by the International Development Research Centre (IDRC) – Canada (grant number: 110025; project number: 26928). We would like to thank Dr. Raphael Nishimura for assisting in study weights computation. We would also like to thank the WOMENA Study Group for supporting this study. We acknowledge BOT (Bridge. Outsource. Transform) for their assistance in data collection crucial for the study’s success.

## Members of the WOMENA Study Group

Jocelyn DeJong, Hala Ghattas, Nisreen Salti, Sasha Fahme, Serena Canaan, Malak Ghezzawi, Hazar Shamas, Ghada E. Saad, Myriam Dagher, Rita Itani, Ali Abboud, and Stephen J. McCall

