## Supplemental Material for "Workplace Violence and Self-Reported Physical and Mental Health: A National Cross-Sectional Study in Lebanon"

### Supplementary Appendix

Figure S1. DAG representing causal relationship between WPV and depressive symptoms

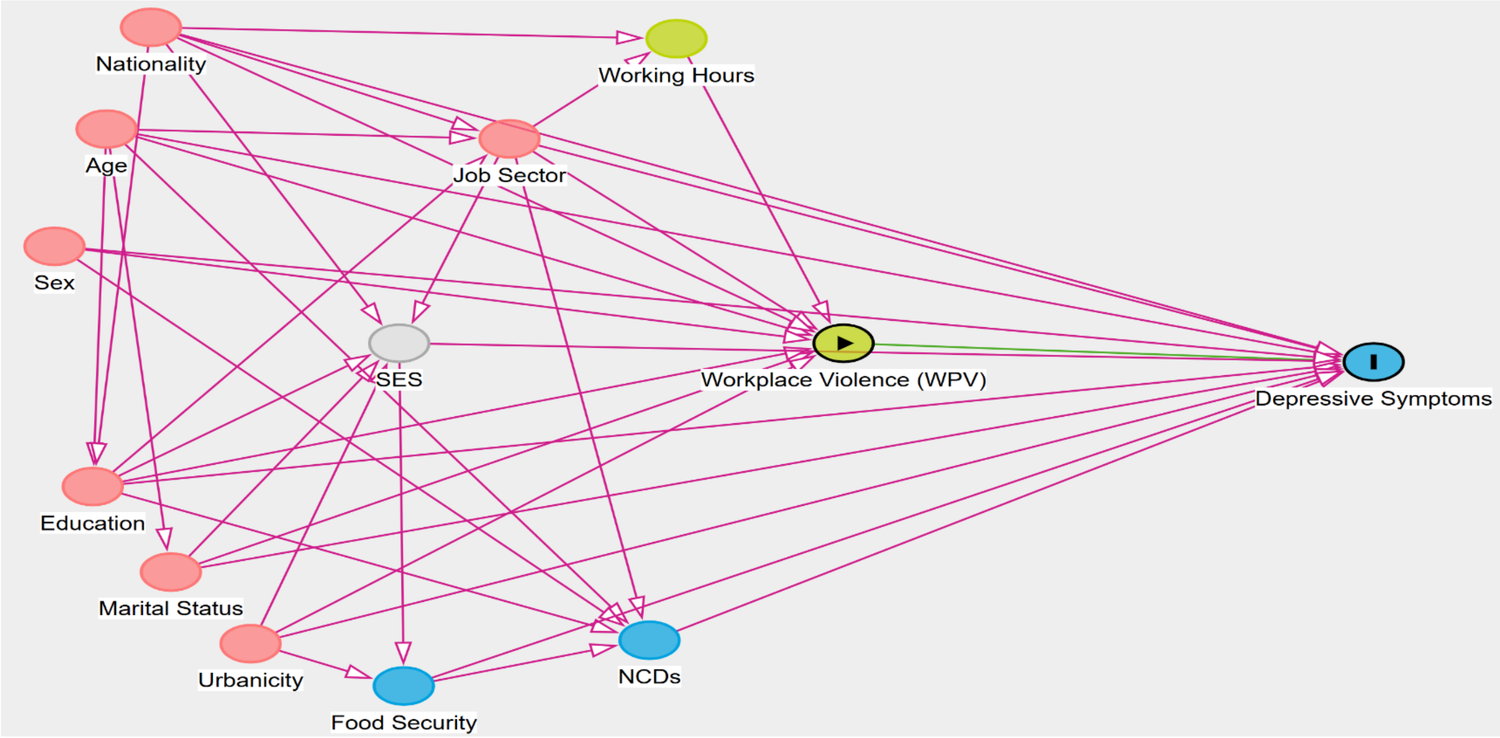

Figure S2. DAG representing causal relationship between WPV and anxiety symptoms

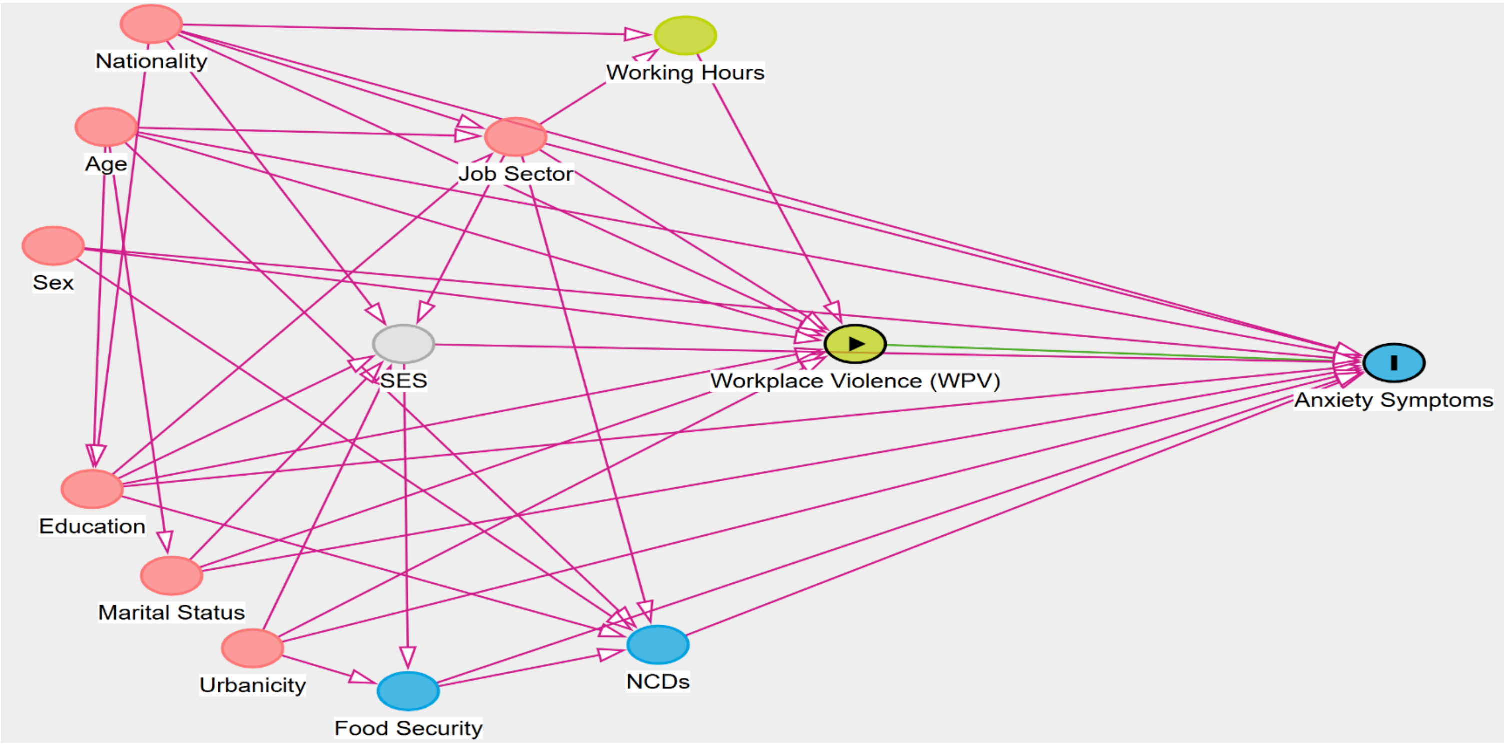

Figure S3. DAG representing causal relationship between WPV and physical health

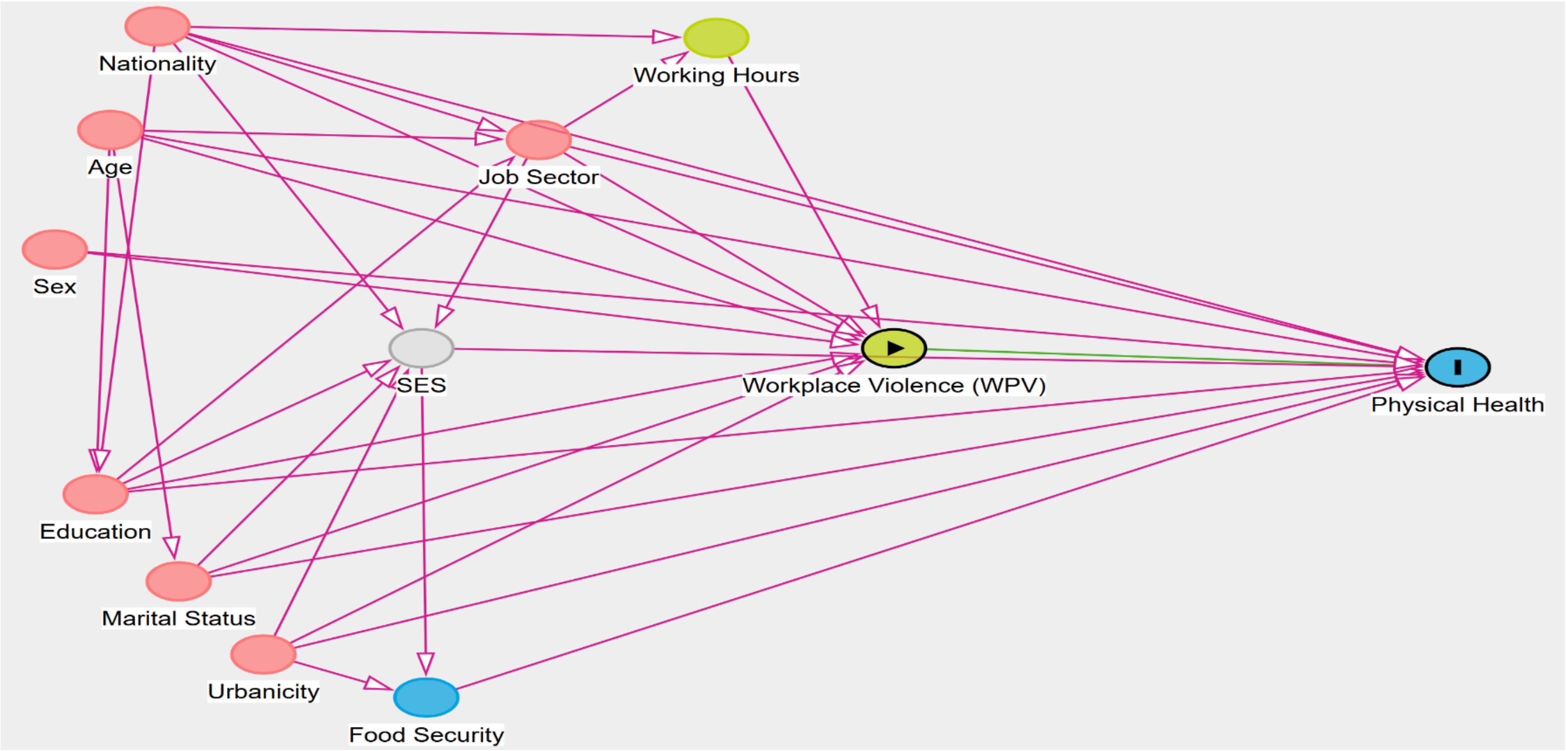

Table S1. Characteristics of participants and their association with depressive symptoms

| | Total | | PHQ9<10 | | PHQ9≥10 | | Unadjusted Odds Ratio (OR) | (95% CI) |
|---|---|---|---|---|---|---|---|---|
| | n=3,036 | (100%) | n= 1,956 | (66·6%) | n= 1,080 | (33·4%) | | |
| **Age median (IQR)** | 37 | (29-47) | 38 | (29-47) | 37 | (29-47) | 0·99 | (0·99-1·01) |
| **Sex** | | | | | | | | |
| Male | 1,865 | (67·4) | 1,222 | (68·2) | 643 | (65·8) | 1·00 | |
| Female | 1,171 | (32·6) | 734 | (31·8) | 437 | (34·2) | 1·11 | (0·95-1·30) |
| **Education** | | | | | | | | |
| No/ Primary Education | 801 | (24·5) | 430 | (20·6) | 371 | (32·3) | 1·00 | |
| Intermediate/Technical | 1,357 | (45·1) | 883 | (45·4) | 474 | (44·5) | 0·62 | (0·51-0·75) |
| College/ Post | 878 | (30·4) | 643 | (34·0) | 235 | (23·3) | 0·43 | (0·35-0·53) |
| **Nationality** | | | | | | | | |
| Lebanese | 1,941 | (75·0) | 1,366 | (79·0) | 575 | (66·1) | 1·00 | |
| Non-Lebanese | 1,095 | (25·0) | 590 | (21·0) | 505 | (33·9) | 1·99 | (1·70-2·32) |
| **Marital Status** | | | | | | | | |
| Single | 721 | (25·1) | 516 | (27·4) | 205 | (20·6) | 1·00 | |
| Married/ Engaged | 2,105 | (68·7) | 1,325 | (67·2) | 780 | (71·6) | 1·41 | (1·17-1·71) |
| Widowed/ Divorced | 210 | (6·2) | 115 | (5·4) | 95 | (7·8) | 1·92 | (1·38-2·67) |
| **Urbanicity** | | | | | | | | |
| Rural | 2,336 | (79·2) | 1,499 | (78·6) | 837 | (80·4) | 1·00 | |
| Urban | 664 | (20·8) | 433 | (21·4) | 231 | (19·6) | 0·89 | (0·73-1·07) |
| Missing | 36 | | 24 | | 12 | | | |
| **Job Sector** | | | | | | | | |
| Government-NGO | 392 | (14·4) | 276 | (15·4) | 116 | (12·5) | 1·00 | |
| Private business | 1,536 | (51·0) | 1,037 | (53·2) | 499 | (46·6) | 1·07 | (0·84-1·38) |
| Private household | 125 | (3·3) | 68 | (2·7) | 57 | (4·6) | 2·12 | (1·37-3·27) |
| Freelance | 977 | (31·3) | 570 | (28·7) | 407 | (36·4) | 1·56 | (1·20-2·02) |
| Missing | 6 | | 5 | | 1 | | | |

Table S2. Characteristics of participants and their association with anxiety symptoms

| | Total | | GAD7<10 | | GAD7≥10 | | Unadjusted OR | (95% CI) |
|---|---|---|---|---|---|---|---|---|
| | n=3,053 | (100%) | n=2,228 | (74.5%) | n=825 | (25.5%) | | |
| **Age median (IQR)** | 38 | (29-47) | 37 | (28-47) | 38 | (29-47) | 1·01 | (0·99-1·02) |
| **Sex** | | | | | | | | |
| Male | 1,874 | (67·3) | 1,403 | (68·4) | 471 | (63·9) | 1·00 | |
| Female | 1,179 | (32·7) | 825 | (31·6) | 354 | (36·1) | 1·20 | (1·03-1·44) |
| **Education** | | | | | | | | |
| No/ Primary Education | 806 | (24·5) | 532 | (22·4) | 274 | (30·8) | 1.00 | |
| Intermediate/Technical | 1,364 | (45·1) | 981 | (44·2) | 383 | (47·6) | 0.78 | (0·64-0·95) |
| College/ Post | 883 | (30·4) | 715 | (33·4) | 168 | (21·6) | 0.47 | (0·37-0·59) |
| **Nationality** | | | | | | | | |
| Lebanese | 1,955 | (75·0) | 1,499 | (77·5) | 456 | (68·2) | 1·00 | |
| Non-Lebanese | 1,098 | (25·0) | 729 | (22·5) | 369 | (31·8) | 1·60 | (1·36-1·89) |
| **Marital Status** | | | | | | | | |
| Single | 731 | (25·3) | 577 | (27·0) | 154 | (20·2) | 1·00 | |
| Married/ Engaged | 2,113 | (68·5) | 1,518 | (67·4) | 595 | (71·7) | 1·41 | (1·14-1·74) |
| Widowed/ Divorced | 209 | (6·2) | 133 | (5·6) | 76 | (8·1) | 1·93 | (1·36-2·74) |
| **Urbanicity** | | | | | | | | |
| Rural | 2,350 | (79·2) | 1,716 | (79·1) | 634 | (79·2) | 1·00 | |
| Urban | 667 | (20·8) | 484 | (20·9) | 183 | (20·8) | 0·99 | (0·81-1·21) |
| Missing | 36 | | 28 | | 8 | | | |
| **Job Sector** | | | | | | | | |
| Government-NGO | 397 | (14·5) | 298 | (14·7) | 99 | (13·8) | 1·00 | |
| Private business | 1,540 | (50·9) | 1,173 | (53·1) | 367 | (44·3) | 0·89 | (0·68-1·16) |
| Private household | 125 | (3·3) | 89 | (3·1) | 36 | (3·7) | 1·28 | (0·80-2·06) |
| Freelance | 985 | (31·3) | 663 | (29·0) | 322 | (38·2) | 1·41 | (1·07-1·84) |
| Missing | 6 | | 5 | | 1 | | | |

Table S3. Characteristics of participants and their association with self-reported poor physical health

| | Total | | Good Physical Health | | Poor Physical Health | | Unadjusted OR | (95% CI) |
|---|---|---|---|---|---|---|---|---|
| | n=3,047 | (100%) | n=673 | (24.3%) | n=2,374 | (75.7%) | | |
| **Age median (IQR)** | 38 | (29-47) | 35 | (27-45) | 38 | (30-47) | 1·01 | (1·00-1·02) |
| **Sex** | | | | | | | | |
| Male | 1,872 | (67·3) | 510 | (79·7) | 1,362 | (63·4) | 1 | |
| Female | 1,175 | (32·7) | 163 | (20·3) | 1,012 | (36·6) | 2·27 | (1·86-2·76) |
| **Education** | | | | | | | | |
| No/ Primary Education | 798 | (24·2) | 132 | (17·5) | 666 | (26·4) | 1 | |
| Intermediate/Technical | 1,365 | (45·2) | 300 | (44·8) | 1,065 | (45·3) | 0·67 | (0·53-0·85) |
| College/ Post | 884 | (30·6) | 241 | (37·7) | 643 | (28·3) | 0·49 | (0·38-0·63) |
| **Nationality** | | | | | | | | |
| Lebanese | 1941 | (74·8) | 490 | (81·0) | 1,451 | (72·6) | 1 | |
| Non-Lebanese | 1106 | (25·2) | 183 | (19·0) | 923 | (27·4) | 1·68 | (1·39-2·04) |
| **Marital Status** | | | | | | | | |
| Single | 727 | (25·2) | 233 | (34·8) | 494 | (22·1) | 1 | |
| Married/ Engaged | 2,112 | (68·6) | 411 | (61·1) | 1,701 | (71·1) | 1·83 | (1·50-2·22) |
| Widowed/ Divorced | 208 | (6·2) | 29 | (4·1) | 179 | (6·8) | 2·67 | (1·69-4·06) |
| **Urbanicity** | | | | | | | | |
| Rural | 2,346 | (79·1) | 513 | (78·1) | 1,833 | (79·4) | 1 | |
| Urban | 668 | (20·9) | 153 | (21·9) | 515 | (20·6) | 0·92 | (0·74-1·14) |
| Missing | 33 | | 7 | | 26 | | | |
| **Job Sector** | | | | | | | | |
| Government-NGO | 396 | (14·6) | 102 | (17·1) | 294 | (13·8) | 1 | |
| Private business | 1,538 | (50·9) | 382 | (56·6) | 1,156 | (49·0) | 1·07 | (0·83-1·39) |
| Private household | 124 | (3·2) | 12 | (1·3) | 112 | (3·9) | 3·6 | (1·85-7·00) |
| Freelance | 983 | (31·3) | 175 | (25·0) | 808 | (33·3) | 1·65 | (1·24-2·20) |
| Missing | 6 | | 2 | | 4 | | | |
